# Weighted-stochastic Avalanche Transition Matrix (ws-ATM): a tool to investigate brain dynamic and its neuropathological alterations

**DOI:** 10.64898/2026.01.08.26343631

**Authors:** Camilla Mannino, Pierpaolo Sorrentino, Marianna Angiolelli, Matteo Demuru, Francesca Trojsi, Mario Chavez, Marie-Constance Corsi

## Abstract

**Background and Objectives:** Amyotrophic lateral sclerosis (ALS) is a progressive neurodegenerative disorder that, beyond motor neuron loss, involves distributed cortical network dysfunction and marked clinical heterogeneity, motivating biologically grounded markers to track disease-related network disruption. Neuronal avalanches provide a framework to probe nonstationary propagation. We introduced a novel functional connectivity metric derived from neuronal avalanches and adapted from the original avalanche transition matrix (ATM) by incorporating activation and avalanche duration: weighted-stochastic ATM (ws-ATM). We hypothesized that embedding temporal persistence would improve robustness and sensitivity to ALS-related alterations, quantitatively linked to standardized clinical severity and staging.

**Methods:** We tested this hypothesis on resting-state MEG source-reconstructed data from 39 individuals with ALS and matched healthy controls. For each neuronal avalanche, we constructed a transition count matrix T, where each element T_ij_ was incremented whenever region i was active at time t and region j at time t+1, thus capturing persistence through consecutive co-activations. Each matrix was then row-normalized to produce a row-stochastic transition probability matrix. Finally, these matrices were aggregated into a subject-level ws-ATM, with weights scaled according to the duration of each avalanche. Robustness was assessed by progressively removing avalanches and quantifying deviations from the full-signal ws-ATM. For clinical relevance, we propose a concordance framework that (i) identifies large-scale reorganization in ALS vs controls, (ii) tests whether individual propagation differences correlate with impairment (ALSFRS-R/MiToS), and (iii) highlights edges whose between-group changes align with within-patient severity in a directionally consistent, clinically interpretable way.

**Results:** ws-ATM converged earlier and stayed stable under substantial avalanche removal, with reduced inter-subject dispersion versus ATM; effects persisted across most truncation levels (up to ∼65% removal). In ALS, ws-ATM detected more altered edges than ATM (370 vs 74), revealing widespread changes with prominent frontal and fronto-motor involvement. Clinical associations strengthened, with greater overlap between ALS–control edges and disability-related edges (ws-ATM: 17/15 for ALSFRS-R/MiToS vs ATM: 6/2), enriching frontal/fronto-motor nodes including superior frontal and precentral regions.

**Discussion:** Our findings indicate that ws-ATM remains reliable with shorter recordings and after removing artifact-contaminated segments, an advantage for clinical diagnostic use. Moreover, it appears to better capture disease-relevant network alterations, providing a biologically grounded set of features that could support ALS stratification.

## Introduction

Amyotrophic lateral sclerosis (ALS) is a progressive neurodegenerative disease characterized by the degeneration of upper and lower motor neurons in the motor cortex, brainstem, and spinal cord, leading to progressive weakness, loss of motor autonomy, bulbar and respiratory failure, and death typically within 3–5 years^1^. It is the most common form of motor neuron disease but is clinically and biologically heterogeneous. The global incidence is low but non-negligible (∼1–3 new cases per 100,000 person-years), with marked geographic variability^2^. Disease severity is usually assessed using the ALS Functional Rating Scale–Revised (ALSFRS-R), which quantifies loss of autonomy in everyday activities across bulbar, motor, and respiratory domains^3^, and the Milano–Torino staging (MiToS) system, which derives discrete disease stages from ALSFRS-R subscores^4^. ALS is increasingly recognized as a diffuse disorder rather than a focal degeneration confined to corticospinal neurons^5^. Structural MRI studies have shown not only cortical thinning of the precentral gyrus and damage to corticospinal tracts, but also involvement of callosal pathways and altered interhemispheric coupling^6^, supporting the view that ALS arises from pathogenic processes affecting the whole brain. Clinically, up to 50% of patients develop cognitive and/or behavioural impairment, and about 13% meet criteria for the behavioural variant of frontotemporal dementia.

Functional connectivity (FC), defined as the statistical dependence between spatially distinct neural time series, has been extensively investigated in ALS using functional MRI (fMRI) and electro/magnetoencephalography (EEG/MEG). These studies demonstrate connectivity changes that extend beyond the primary motor cortex to large-scale frontoparietal and frontotemporal networks, including patterns that converge toward a frontotemporal-dementia–like organization in patients with cognitive or behavioural involvement^7^. MEG studies have further revealed large-scale alterations in oscillatory activity and connectivity, implicating frontal executive and integrative circuits alongside corticospinal output^8,9^. Despite this progress, many FC approaches provide an aggregate description of activity over a predefined time interval, implicitly assuming stationarity. This assumption can be suboptimal given evidence that large-scale brain activity is dynamic and multiscale. Resting activity shows lawful oscillations nested within aperiodic, scale-free bursts of activity^10;11^. Capturing these dynamics may improve sensitivity to subtle, spatially distributed pathological changes. A prominent dynamical framework describes resting activity in terms of neuronal avalanches: cascades of burst events that spread through networks.

In a large resting-state MEG sample, Shriki and colleagues showed that spontaneous cortical activity organizes into avalanches rather than random fluctuations^12^. Avalanches do not propagate randomly but preferentially follow white-matter bundles that constitute the structural connectome^13^, resulting in differential recruitment of specific brain regions^14^. Avalanche dynamics have been leveraged in several contexts. In epilepsy, avalanche-based analyses have been used to study alterations in propagation and network recruitment^15,16^. In Brain-Computer Interface (BCI) paradigms, avalanche measures have been proposed as markers of task engagement and learning, and as predictors of subsequent BCI control performance^17,18^. In ALS, previous work suggested that the “functional repertoire” (the number of unique avalanche patterns expressed during a recording) can serve as a non-invasive marker of altered brain dynamics and potentially of disease-related network constraint^20^. Building on these ideas, avalanche transition matrices (ATMs) were introduced as a compact description of propagation. Polverino and colleagues used this framework to show that ALS patients exhibit increased centrality in both cortical and subcortical regions compared with healthy controls, in proportion to disease duration^19^. ATMs have also been explored as alternative feature representations for BCI classification tasks^20,21^.

In this work, we introduce a novel functional connectivity metric derived from neuronal avalanches and adapted from the original ATM. We hypothesize that incorporating temporal information will enhance the metric’s robustness and increase its sensitivity to brain dynamics, particularly those associated with neurological disorders. To test this hypothesis, we apply the novel metric to MEG data from individuals with ALS and matched healthy controls, with the goal of capturing disease-related network disruption in a way that can be quantitatively linked to standardized clinical staging and potentially serve as a biologically grounded biomarker for progression and stratification.

We first evaluate the robustness-related theoretical properties of the proposed weighted-stochastic ATM (ws-ATM) and compare its performance with the original ATM. We then investigate its clinical relevance using two complementary approaches: (i) a group-level analysis to detect ALS-related alterations in brain network dynamics relative to healthy controls, and (ii) a subject-level analysis examining associations between ws-ATM–derived features and clinical severity scales. Finally, we determine which edges and nodes show significant effects consistently across both analyses, where nodes represent brain regions and edges represent the functional connections between pairs of nodes, quantified here by ws-ATM/ATM transition-based coupling.

## Method

### 2.1. Participants

Thirty-nine ALS patients (29 males and 10 females; mean age 59.63 years; SD ± 12.87; mean education 10.38 years; SD ± 4.3) were recruited from the Hermitage Capodimonte Clinic in Naples and collaborating institutions. ALS was diagnosed according to revised El Escorial criteria^22^. Inclusion criteria were absence of major medical illness and substances that could interfere with MEG signals, no other major systemic, psychiatric, or neurological diseases, and no focal or diffuse brain damage reported at MRI assessment. The control group comprised thirty-nine subjects (28 males and 11 females) matched for age (64.0 ± 10.4) and education (12.0 ± 4.3). The ALSFRS-R total score^3^ was used to quantify functional status (higher scores indicate better function). MiToS staging^4^ was used to assess disease stage from ALSFRS-R subscores.. The study protocol was approved by the Local Ethics Committee (University of Campania “Luigi Vanvitelli”, Italy) and all participants provided written informed consent in accordance with the Declaration of Helsinki.

### 2.2 Acquisition methods and preprocessing

Resting-state MEG was recorded in a magnetically shielded room using 154 SQUID magnetometers plus nine reference sensors. Head position was tracked with a Polhemus Fastrack digitizer using four anatomical landmarks and four head coils. Each participant completed two eyes-closed resting runs (3.5 min each) separated by a 1-min break, with simultaneous ECG and EOG for artefact detection. Data were anti-alias filtered and sampled at 1024 Hz. Preprocessing (FieldTrip toolbox^23^ in MATLAB) included 0.5–48 Hz 4th-order Butterworth band-pass filtering, PCA for environmental noise reduction, and ICA to remove physiological artefacts (typically one ECG component; EOG component rarely). After discarding the noisiest segments, runs were concatenated to obtain a denoised, equal-length time series per participant (122.8 s). MEG was co-registered to individual MRI and source-reconstructed to Automated Anatomical Labeling^24^ (AAL)-defined cortical regions. If MRI was unavailable (6 patients, 9 controls), a standard template was used. For further details refer to Sorrentino et al. (2018)^8^. In this work, subcortical and cerebellar AAL regions were excluded due to limited MEG sensitivity^25^, yielding N = 78 cortical regional time series (122.8 s) for each participant.

### 2.3 Neuronal Avalanches detection

Neuronal avalanches were identified in continuous MEG by converting each regional time series into binary events via z-scoring and amplitude thresholding. An avalanche started when at least one brain region exceeded the threshold and ended when all regions returned below threshold. Following our previous work^17,20^, detection depended on two parameters: the z-threshold (θ_av_), defining which suprathreshold activations belong to an avalanche, and a minimum avalanche duration (λ_min_av_), used to discard very brief events likely driven by noise/artifacts *(Figure 1)*. Careful calibration of these parameters is essential to robustly characterize meaningful neurophysiological dynamics while minimizing contamination from non-neural noise.

**Figure 1:**
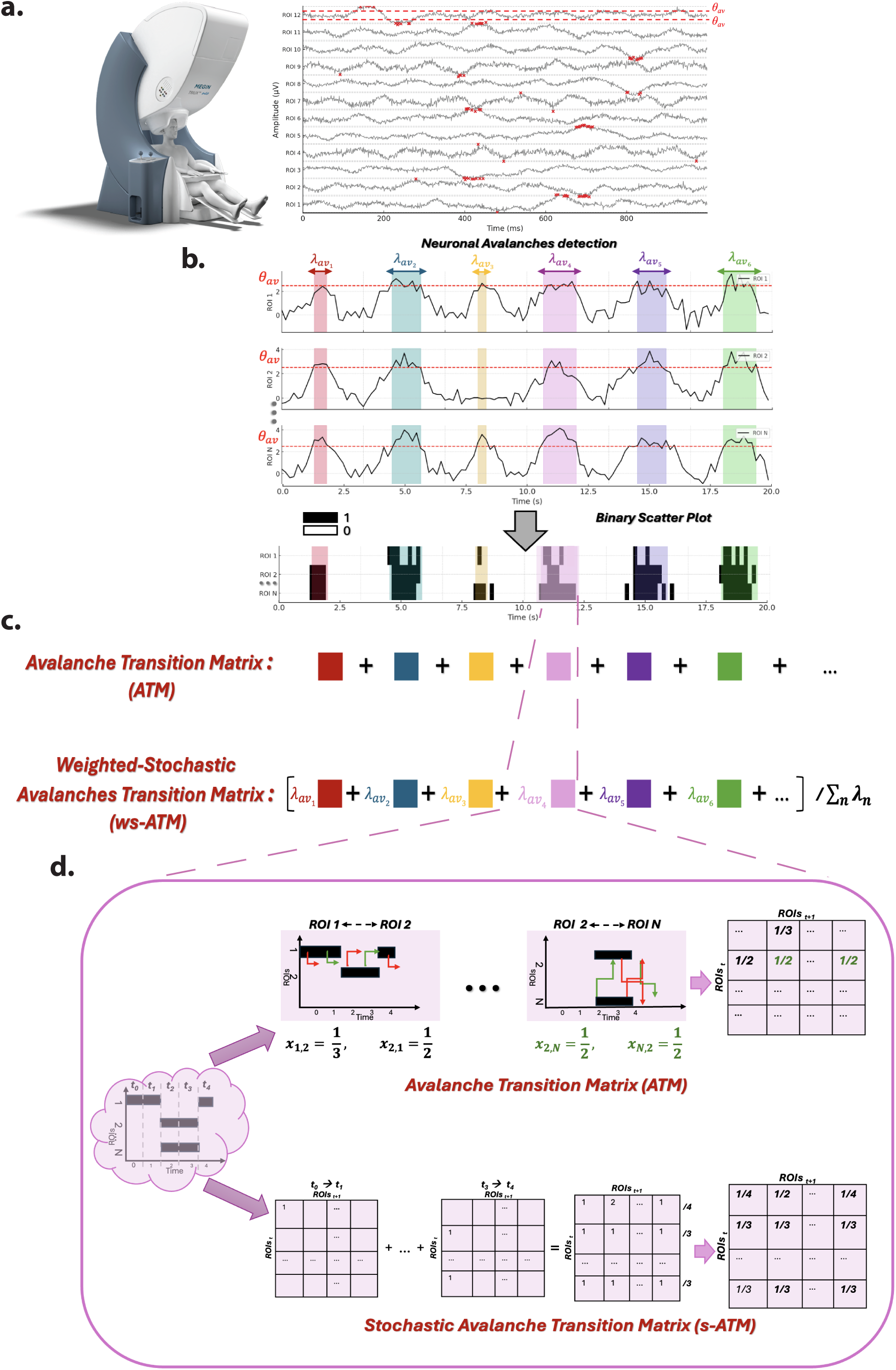
Methodological framework overwiev: a) MEG setup and burst over-threshold brain activity; b) Neuronal Avalanches detection; c) Avalanche Transition Matrix (ATM) and weighted-stochastic Avalanche Transition Matrix (ws-ATM) formulation at subject level, different neuronal avalanches integration; d) focus on one single avalanche matrix: ATM and ws-ATM at avalanche level.

To set θ_av_, we compared each participant’s normalized signal distributions (across subjects and ROIs) with best-fit Gaussians; divergence from Gaussianity was used as a marker of nonlinear, avalanche-like dynamics (as opposed to Gaussian fluctuations expected from summed independent noise sources). Although deviations emerged around ∼2.4σ (CTRL) and ∼2.5σ (ALS), we set a threshold of 4σ, in line with prior work^17^, as a conservative trade-off that maximized separation from Gaussian trends while keeping avalanche counts and durations physiologically plausible and avoiding excessive detections. The selection of λ_min_av_ was informed by established anatomical evidence indicating that interhemispheric transmission delays can be as short as 5 ms for large myelinated callosal fibres^26^. This constraint ensured that the temporal resolution of detected avalanches remained consistent with known neurophysiological conduction delays.

### 2.4 Avalanche transition matrix (ATM)

The Avalanche Transition Matrix (ATM) quantifies how likely neuronal avalanches propagate between pairs of brain regions. In the original ATM, element 𝐴𝑇𝑀_!"_is the probability that region 𝑗 is active at 𝑡 + Δ𝑡 given that region 𝑖 was active at 𝑡 during an avalanche. For each avalanche, a transition matrix is computed, then matrices are averaged elementwise and symmetrized to obtain a subject-specific ATM. This approach summarizes pairwise spreading probabilities but does not model stochasticity or activation duration, collapsing transitions over time without distinguishing short vs. long activations/avalanches and ignoring non-homogeneous sampling *(Figure 1)*. Because brief co-activations are more likely transient or noise-driven, whereas longer co-activations reflect more stable recruitment and stronger coupling (similarly for avalanche length), omitting duration may underweight sustained propagation and overemphasize short, potentially artefactual events. To overcome this, we reformulated the ATM as a weighted-stochastic model that explicitly incorporates transition probabilities together with regional activation periods and avalanche durations.

### 2.5 Weighted-stochastic Avalanche Transition Matrix (ws-ATM)

Building upon the previous formulation, the original ATM (ATM) was further developed into a weighted-stochastic matrix, ws-ATM *(Figure 1)*, designed to capture both probabilistic relationships between ROIs and activation durations, thereby enabling the application of Markov chain^27,28^ modelling to brain network dynamics.

#### Step 1: Transition Counting

For each time point 𝑡, we treat the activity pattern as defining a Markov-like state. We then count time-shifted co-activations: if ROI 𝑖 is active at 𝑡 and ROI 𝑗 is active at 𝑡 + 1 in the binary activity matrix 𝐴(𝑖, 𝑡), we increment the transition count 𝐶(𝑖, 𝑗) *(Figure 1d)*.

The number of consecutive activations between these two ROIs, 𝐶(𝑖, 𝑗), is incremented as:

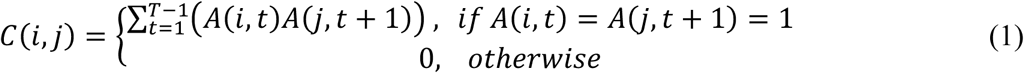

where A(i,t) is the binarized activity of ROI i at time t within each avalanche, and T is the total time.

#### Step 2: Probability Normalization

For each ROI 𝑖, the transition probabilities are obtained as:

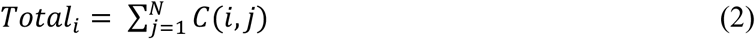

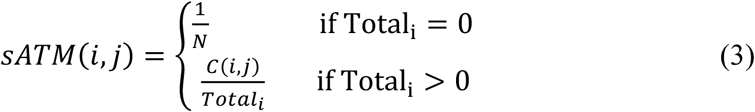

Where N is the total number of ROIs.

If no transitions are observed for a specific ROI (row sum = 0), uniform probabilities are assigned to preserve the stochastic property:

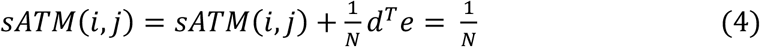

where 𝑒 is a column vector of ones (𝑁 × 1) and 𝑑 is a vector of ones that activates the fallback distribution.

This stochastic formulation allows the ATM to serve as a transition probability matrix in a Markov chain framework. By incorporating the duration of activations, this method provides more detailed information on temporal engagement of ROIs and the strength of connectivity between specific pairs of regions. Consequently, the stochastic ATM (s-ATM) enables a more refined modelling of spatiotemporal brain activation patterns, identifying the most prominently involved ROIs and quantifying both the strength and persistence of their interactions.

#### Step 3: Weighted averaged across avalanches

A stochastic ATM is firstly computed for each individual avalanche. To integrate these matrices into a single subject-level representation, a weighted averaging procedure is then applied *(Figure 1c)*. Specifically, each s-ATM is multiplied by the corresponding avalanche length prior to averaging, thereby assigning greater weight to longer avalanches. This weighting reflects the assumption that longer avalanches are more likely to capture meaningful brain dynamics, whereas shorter avalanches may be more susceptible to noise or artifacts. The final subject-specific ws-ATM is obtained by normalizing the weighted sum across all avalanches.

### 2.6 ATMs Convergence

To investigate the contribution of neural avalanche dynamics to the organization of the functional transition structure, we systematically removed avalanches and quantified their effect on control subjects using both ATM formulations, x-ATM *(Figure 2a)*. By progressively removing avalanches from the entire signal for each subject, we incrementally eliminated segments of the underlying temporal structure, to test for convergence with respect to the overall average (i.e., considering the whole signal). To this end, at each removal level, we recomputed the averaged ATM across the avalanches and quantified the corresponding deviation from the baseline (no-removal) ATM using the L1 edge difference metric, providing a measure of the global alteration in transition structure, as in:

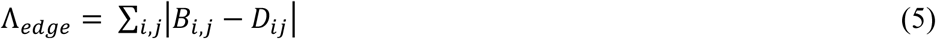

**Figure 2:**
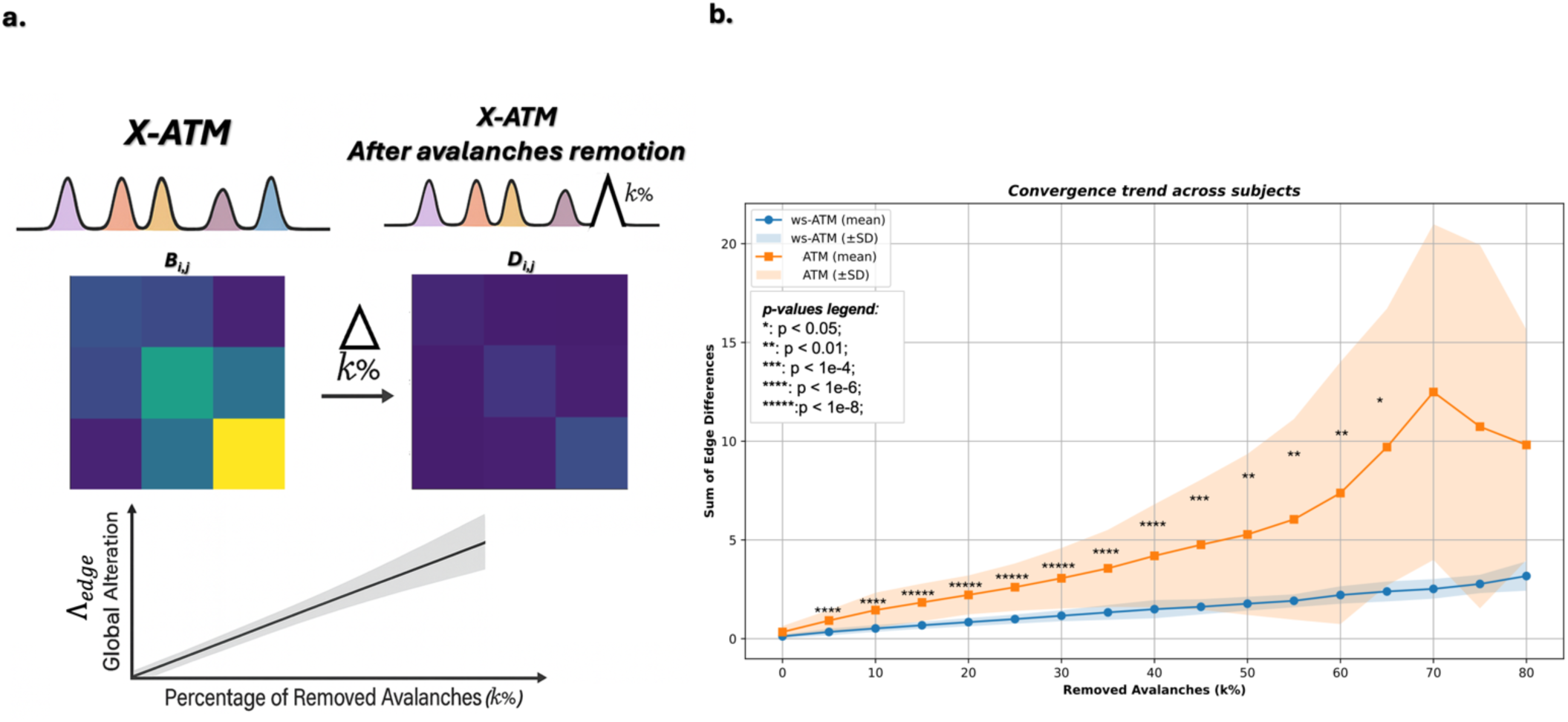
**ATM Convergence: a) Methodological approach, b) Different impact of avalanches removal from the end of the signal across subjects between ATM and ws-ATM in healthy subjects**; Orange curve : Avalanches-Transition Matrix (ATM) trend; Blue curve : weighted-stochastic ATM (ws-ATM) trend

This procedure allows us to characterize the extent to which the network’s transition structure remains robust despite substantial event removal.

Finally, we assessed the different behaviour between the two ATM formulations (x-ATM) utilizing a t-test with FDR correction for multiple comparison for each single percentage of removed avalanches. We first performed this analysis in control subjects and subsequently validated our findings in ALS subjects. To test the robustness of the results in controls, we also repeated the procedure with avalanches removed at random positions within the signals (as opposed to removing the n% of the last avalanches).

### 2.7 Concordance of network alterations and clinical impairment in ALS subjects

To investigate differences in brain network organization between healthy controls and individuals with amyotrophic lateral sclerosis, we performed a group-level statistical analysis on each edge of the avalanche transition matrix (ATM and ws-ATM). Specifically, a two-sample t-test was applied to each individual edge to identify connectivity alterations associated with the disease *(Figure 3a)*. In a second step, to identify potential biomarkers of the clinical severity, we examined the relationship between edge weights and clinical severity by computing Spearman’s rank correlation coefficients with scores on the ALSFRS-R^3^ (and the MiTos staging^4^ system, reported in the Supplementary Materials).

**Figure 3:**
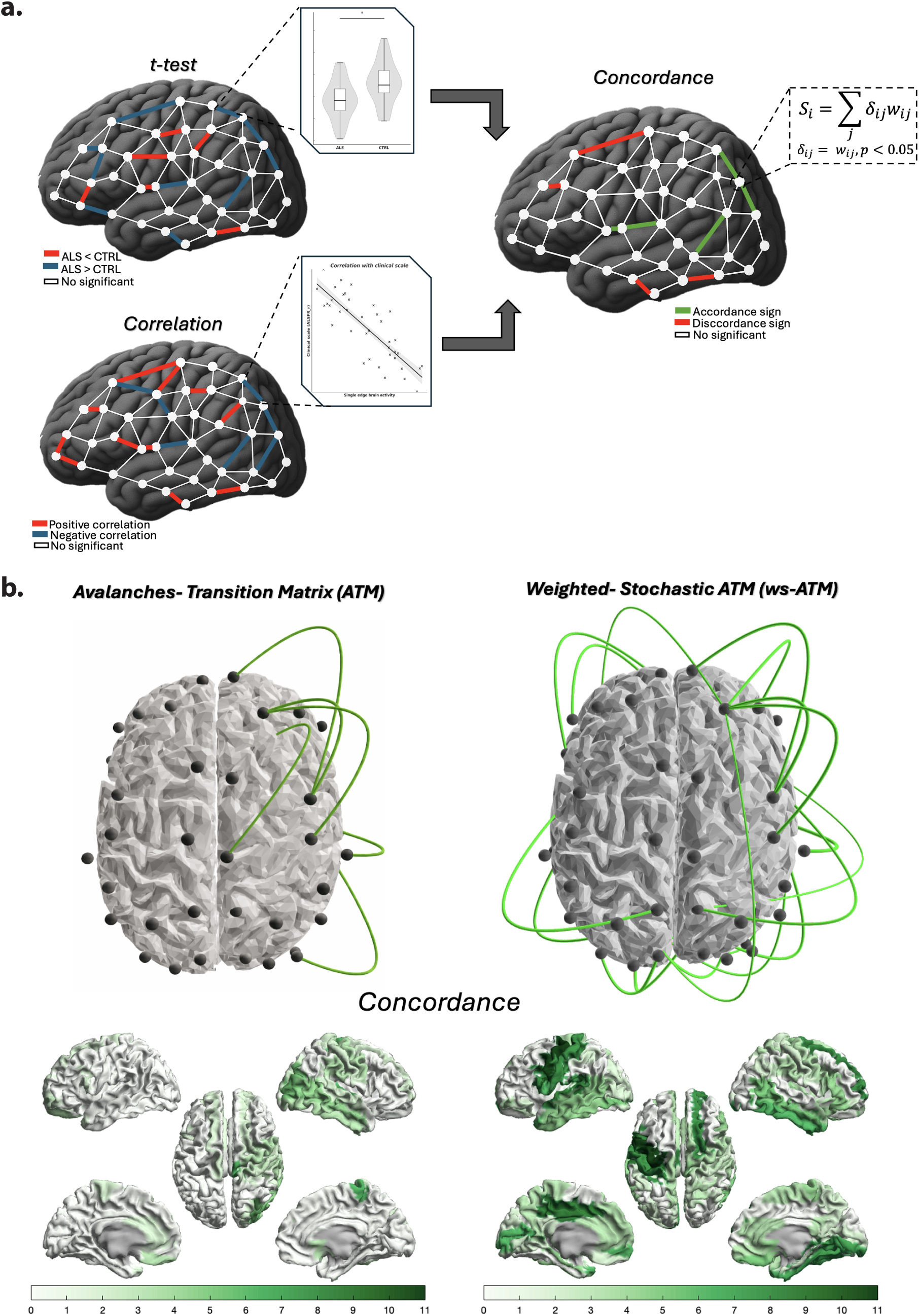
Clinical validation framework: a) **Framework explanation:** (i) a **group-level comparison** between ALS patients and healthy controls, (ii) a **single-subject correlation analysis** between neuronal avalanche dynamics and clinical disability (ALSFRS-R), and (iii) a **concordance validation**, which examines the consistency between disease-related network alterations and clinical progression b) **Concordance t-test and correlation with clinical scale results using ATM and ws-ATM at edges level and at nodal level.** Edges with concordant sign are in green, edges with discordant sign are in red. All counts refer to edge-level tests at p < 0.05 without multiple-comparison correction.

To evaluate the concordance of network alterations, differences observed in ALS subjects compared to the controls, with disease severity, we first assessed the overlap between edges showing significant group differences and those significantly correlated with clinical scores. Specifically, we retained edges that differed between groups (t-test, p < 0.05) and edges whose weights correlated with clinical measures (Spearman, p < 0.05), and then examined whether the direction of these network perturbations (i.e., the sign of edge differences relative to controls) was consistent with the pattern observed across disease severity *(Figure 3a)*.

To facilitate interpretation, at each step, we computed the number of significant edges summed across each column of the ATMs, providing a region-wise summary of network alterations *(Figure 3a)*. This topological measure reflects the overall involvement or influence of each node within the network and allows us to interpret clinically relevant differences in term of regional alterations.

## Results

### 3.1 Increase robustness and stability using weighted-stochastic ATM

Progressively truncating the last avalanches of each trial highlighted a marked difference in the stability between the ws-ATM and the original ATM in controls subjects. In ws-ATM formulation the error introduced by the removal increased only shallowly and near-linearly, remaining essentially unchanged through ∼50% removal, which shows fast convergence *(Figure 2b blue line)*. By contrast, the error introduced by the removal in the ATM rose steadily; deviations escalated sharply, and across-subject variability widened with a slower convergence *(Figure 2b orange line)*. Group-level comparisons across subjects validated these patterns. The FDR-adjusted p-values (q-values) were extremely small for removal proportions ranging from 1% to approximately 60% (typically q-values < 10⁻⁶ and remaining below 5 × 10⁻³ up to ∼58%), demonstrating a robust separation between ws-ATM and ATM across most of the truncation range. The maximum statistical difference between the two formulations was observed for removal proportions below 30%, where q-values reached values lower than 5 × 10⁻⁹. Statistical significance was maintained up to approximately 64–65% removal but was lost beyond 66%, coinciding with a marked increase in variance for the ATM formulation, which inflated standard errors and reduced statistical power despite larger mean differences.

A very similar pattern emerged when avalanches were removed at random positions in the time series rather than only from the end *(Figure 2b; Supplementary Figure 2a)*, and an analogous trend was observed in ALS patients, where ws-ATM showed reduced inter-subject variability and a slower change in edge differences with increasing avalanche removal compared with original ATM algorithm *(Supplementary Materials, Figure 2b)*. Taken together, these findings indicate that later avalanches add little unique transition information to ws-ATM, whereas ATM depends disproportionately on them and becomes unstable when they are removed. Consequently, ws-ATM is more sample-efficient, remains stable when avalanches are removed, and yields more reproducible results across subjects.

### 3.2 Edges-identifiability: ALS

After establishing the stability of ws-ATM in control participants, we subsequently examined whether avalanche-based connectivity alterations derived from ws-ATM in ALS patients carry clinically meaningful information. Specifically, we assessed whether this approach discriminates ALS patients from healthy individuals, captures relationships between network alterations and disability severity, and yields patterns consistent with current neurophysiological understanding of the disease. Comparing healthy subjects with ALS revealed widespread alterations in the transition matrix. Using the original ATM, we detected 74 edges (i.e. elements of the x-ATM matrix) with significant group differences (p<0.05, uncorrected), primarily temporal– occipital links, consistently observing higher transition probabilities in ALS as compared to healthy controls *(Supplementary Materials, Figure 3)*. The analysis of ws-ATM led to the identification of 370 significant edges, distributed across intra-and inter-lobe connections among frontal, parietal, temporal and occipital cortices focus predominantly on the frontal lobe *(Supplementary Materials, Figure 3a)*. In addition to that, homogeneous distribution of positive and negative brain alteration across the different brain regions is observable in patient data compared to the healthy one, showing hyper-connection and segregation phenomena.

We next intersected edges that differed across conditions with those showing significant associations with clinical severity. The majority of significantly correlated edges is detected in frontal lobe (a two-fold increase with respect to the other lobes) using both transition matrix formulations *(Supplementary Materials, Figure 3)*. For ATM, significant correlations with ALSFRS-R were mainly negative. In contrast, ws-ATM yielded a more mixed pattern, with a larger share of positive correlations and fewer, more spatially scattered negative associations. An analogous pattern with opposite sign was observed for MiToS, as expected for a scale that increases with disease severity *(Supplementary Materials, Figure 4).* The overlap between the two analyses was modest for ATM but substantially larger for ws-ATM (ATM: only 6 and 2 edges for ALSFRS and MiToS, respectively; ws-ATM: 17 and 15, respectively *(Supplementary Materials, Figure 3-4)*, with a predominance of clinically coherent edge directions *(Figure 3b)*. Correlations with ALSFRS revealed complete concordance in the direction of improvement between the ALS–CTRL contrast and the clinical correlation *(Figure 3b)*. In MiTos, edges indicating improvement in the same direction *(green connectome edges)* were predominant, whereas incoherent edges *(red connectome edges)* were fewer and spatially scattered *(Supplementary Materials, Figure 4)*. These results are encouraging, as we interpret concordant edges as more likely to carry genuine disease-related information, while the few scattered incoherent edges are more plausibly attributable to noise.

The distribution of edges per lobe meeting both criteria was consistent across both transition-matrix formulations, peaking in the frontal lobe; notably, with ws-ATM, we could identify frontal regions where the activities seem to difference in a way proportional to clinical disability, such as the Superior Frontal cortex and motor areas including the Precentral ROIs *(Figure 3b)*. Overall, ws-ATM not only increases sensitivity to ALS-related network disruptions but also enriches for edges whose directional changes align with clinical status, thereby strengthening the interpretability of the pathological connectome signature.

## Discussion

### 4.1 Weighted-stochastic Avalanche Transition Matrix proprieties and advantages

The ws-ATM extends the original avalanche-based connectivity framework by incorporating a probabilistic, temporally weighted formulation of neuronal propagation. Within this model, each matrix element represents the likelihood that activity in one cortical region temporally follows activity in another. *(Figure 1c)*. This formulation embeds avalanche dynamics, allowing the computation of the connectivity matrix for every new detected event. Moreover, this structure enabling the use of stochastic-process theory to characterize brain network evolution over time perspective, establishing, also, a direct link with brain network proprieties and tools from graph theory^29,30^. For example, the PageRank application is now applicable, a based centrality measures, where the stationary distribution of the transition matrix identifies brain regions with high persistence or influence within the functional network.

Within this probabilistic structure, ws-ATM shows improved reliability compared with the original ATM. By weighting transitions according to activation duration and normalizing them stochastically, ws-ATM down-weights brief, noise-driven avalanches. As a result, it displays reduced inter-subject variability and a more homogeneous description of cortical propagation *(Figure 2b; Supplementary Materials, Figure 2)*, which is particularly important in heterogeneous clinical populations and in noisy, real-world scenarios. The ws-ATM is also more robust to data loss: when avalanches are progressively removed, it converges early and deviates only modestly from the full model. This indicates that ws-ATM rapidly captures the essential transition structure and remains stable. In contrast, the original ATM exhibits late convergence, larger dispersion, and a stronger dependence on the inclusion order and number of avalanches, making it more sensitive to sampling variability and to artifact removal. Consequently, ws-ATM supports the reliable estimation from shorter recordings and tolerates more aggressive removal of noisy segments without substantially degrading the estimates, offering clear advantages for clinical applications. For example, MEG recordings frequently contain eye blinks and muscle artifacts, a problem that is exacerbated in patients with motor disorders who may present dystonia, spasticity, or difficulty maintaining a stable seated position over prolonged periods.

### 4.2 Clinical validation

In this work, we present a framework that embeds the weighted-stochastic Avalanche Transition Matrix into a multi-step pipeline to characterize both global and individual alterations in brain dynamics in ALS. First, we use this framework to compare avalanche-based connectivity between ALS patients and healthy controls, identifying large-scale patterns of network reorganization at the group level. We then move to a single-subject perspective, correlating both ATM measures with clinical disability (ALSFRS-R) to test whether individual differences in avalanche propagation are systematically related to functional impairment. Finally, we perform a concordance analysis that jointly inspects the direction and spatial distribution of ALS–control differences and clinical correlations, asking whether edges that change between groups also vary with disease severity in a clinically coherent way. Taken together, these steps define an integrated framework *(Figure 3a)* that links avalanche-based network alterations, to both between-group contrasts and within-patient clinical progression, providing a structured way to evaluate the clinical relevance of avalanche-based dynamics. By combining these complementary analyses, our approach offers a comprehensive and clinically grounded characterization of avalanche propagation in ALS, identifying the cortical hubs most affected by the disease and contextualizing their reorganization within the continuum of neurodegeneration. This methodology extends prior evidence of altered and spatially constrained brain dynamics across neurodegenerative disorders^18,19,31,32,^, demonstrating how a dynamic-systems perspective can reveal both compensatory and pathological aspects of network reorganization in ALS.

When comparing ALS patients with healthy controls, ws-ATM revealed widespread alterations in brain dynamics across multiple cortical regions, characterized by the coexistence of abnormally increased coupling, hyperconnectivity, and reduced integration, functional segregation, within the same network. In the supplementary figures, these two patterns are visualized as areas of excessive synchronization versus relative disconnection from the global network *(Supplementary Materials, Figure 3a)*. In contrast to the original ATM formulation, which tend to detect an hyperconnectivity, ws-ATM offers a more comprehensive picture of ALS-related network pathology. This dual pattern mirrors prior neuroimaging observations that ALS involves the coexistence of increased connectivity and progressive interhemispheric segregation^6,33,34^. Agosta et al. (2011)^33^ reported elevated resting-state coupling between the primary sensorimotor cortex and contralateral associative and cerebellar regions, likely reflecting early compensatory recruitment. Conversely, Verstraete et al. (2010)^6^ and Schmidt et al. (2014)^34^ demonstrated reduced interhemispheric integration within motor and premotor cortices, pointing to structural disconnection and network fragmentation.

Within this framework, the correlation analyses with clinical impairment further emphasize how hyperconnectivity and isolation coexist dynamically, representing complementary expressions of ALS progression. The two ATM formulations revealed different correlation trends with clinical disability. In the ATM, most significant edges showed negative correlations with ALSFRS-R, suggesting that increased connectivity was associated with greater disability, potentially reflecting maladaptive hyper-synchronization and reduced network efficiency. Conversely, the ws-ATM exhibit also an increasing number of positive correlations, implying that the disease’s progression should be associated with the isolation of specific brain regions from the central brain network *(Supplementary Materials, Figure 4a)*. This difference might stem from the distinct weighting of short and long neuronal avalanches in the two models, indeed ws-ATM emphasizing longer propagation sequences than ATM. The divergence between hyperconnectivity and segregation also reflects the frequency-band sensitivities of the two formulations. Higher-frequency oscillations are associated with longer-lasting avalanches that propagate across broader cortical brain regions^35^. In contrast, lower-frequency oscillations giving rise to shorter, spatially constrained cascades^36^. Moreover, previous studies have shown that γ-band activity exhibits an inverse trend relative to low-frequency dynamics during ALS progression^7,37,8^. Elucidating the mechanistic relationship between avalanche duration and the underlying oscillatory processes, however, remains an open question and lies beyond the scope of the present work.

Both ATM models consistently identified the frontal and motor cortices, particularly the precentral gyrus and right superior frontal gyrus, as key hubs. However, the ws-ATM highlighted a somewhat different profile, with more significant edges in the left anterior cingulate cortex, left para-hippocampal gyrus, and bilateral temporal poles. Although these regions fall outside the principal motor-frontal hub, they are part of deep and limbic circuits implicated in ALS progression, particularly those associated with memory and emotional regulation. The identified frontal and motor regions align with previous evidence. Sorrentino et al. (2018)^8^ reported disease-related activity in the right middle frontal gyrus, right precentral gyrus, and right pallidum, all of which correlated with disease duration. These areas are recognized as core nodes of ALS pathophysiology, encompassing both motor and extra-motor cortical regions. Notably, previous analyses based on nodal strength to decode the neuronal avalanche spreading in ALS did not identify significant correlations with clinical scales such as King’s, MiToS, or ALSFRS-R. In contrast, our study identified multiple edges significantly correlated with both ALSFRS-R and MiToS scores *(Supplementary Materials, Figure 3-4)*, across both ATM formulations. This confirms that a posteriori edge-count mapping employed here yields a more data-driven overview of cortical regions implicated in ALS, avoiding strong a priori constraints and capturing subtle yet meaningful dynamic alterations.

The final stage of our analysis assessed the consistency between connectivity abnormalities identified in ALS patients and their associations with disease progression, thereby providing cross-validation of the two analytical approaches. Specifically, we examined whether group-level differences between patients and controls occurred along the same network edges that, within the patient group, exhibited proportional relationships with individual disability severity. These analyses were conducted independently, yet revealed a substantial overlap, strongly indicating that ws-ATM captures pathophysiologically meaningful processes. Moreover, the directionality of the group differences was concordant with the observed within-patient associations: edges showing increased values in patients relative to controls were characterized by positive correlations with disability, whereas edges exhibiting reduced values in patients showed inverse correlations.

We interpret these findings as evidence of a relationship between clinical impairment and functional network reorganization. This concordance was observed across a greater number of significant edges in ws-ATM compared to the original formulation, highlighting its enhanced sensitivity to spatially distributed reorganization in ALS. Both models consistently identified the frontal and motor cortices as key network hubs *(Figure 3b).* However, the conventional ATM exhibited a right-hemispheric predominance, consistent with previous reports of increased functional connectivity in the right precentral and superior frontal gyri, the fronto-parietal network, the angular gyrus, and the inferior cerebellum^38,39^. This right-lateralized hyperconnectivity likely represents a compensatory mechanism for the left-predominant neurodegeneration typical of ALS^40,41^. The right frontal hubs identified here mirror those most commonly affected on the left side in ALS pathology^42,43,44^.

Taken together, these findings indicate that the ws-ATM provides a more complete and neurophysiologically meaningful representation of ALS network reorganization, capturing both bilateral motor involvement—linked to motor impairment—and right frontal network recruitment, associated with the cognitive decline frequently observed in ALS^45,46,47^. The middle frontal and precentral gyri form part of the motor network and are known to undergo progressive grey- and white-matter degeneration as the disease advances^48,49^. Supporting these observations, Borgheai et al. (2020)^50^ demonstrated a shift toward a more centralized frontal functional network in ALS, identifying high-degree hubs in the right prefrontal cortex. Together, these findings reinforce the view of ALS as entailing a network-wise degeneration^51^, circuit-specific vulnerability^52^, and disease propagation along structural connectivity pathways^6^. The same ideas are now being corroborated also by evidence from neuroimaging, neuropsychology, and post-mortem analyses^53,54^.

During progressive data truncation, ws-ATM consistently captured clinically relevant differences, identifying approximately three- to five-fold more significant edges than ATM. This held true both for all significant connections and for concordant edges, defined as connections where ALS–control differences aligned with correlations to clinical scores. This advantage was particularly pronounced in frontal and motor cortices. In contrast, ATM identified a smaller but highly concordant ALSFRS-R network, which began losing its fronto-motor component after removing roughly 25–30% of avalanches, indicating rapid loss of sensitivity to clinically relevant hubs with reduced data length. By comparison, ws-ATM exhibited a more robust fronto-motor organization: frontal and motor ROIs remained significant even when up to 70–75% of avalanches were removed, with their disappearance occurring only after an overall ∼60% reduction in significant edges (data not shown).

### 4.3 Limitations & future directions

Despite these promising results, several limitations should be acknowledged. First, subcortical regions were not included in the present analysis due to the intrinsic limitations of MEG in detecting deep sources. Nevertheless, subcortical structures such as the basal ganglia^55,56^ and the pallidum likely carry clinically-relevant information. Incorporating subcortical ROIs through multimodal approaches, such as combined MEG–MRI or EEG–fMRI analyses, could therefore provide a more comprehensive characterization of whole-brain avalanche dynamics and their alterations in ALS. In addition, prior work has shown that neuronal avalanches can be reliably captured using EEG in healthy individuals, with results comparable to those obtained from MEG recordings^20^. Accordingly, future studies should study scalp EEG data to establish a more accessible and efficient platform for both clinical and research applications.

Secondly, this study employed a cross-sectional design. Longitudinal recordings in the same individuals across different disease stages would allow tracking of disease progression within subjects, reducing inter-individual variability and enabling personalized modelling of disease trajectories. Recent work has shown that neuronal avalanche dynamics can predict individual behavioural performance in brain–computer interface tasks^17^, supporting the notion that these dynamics reflect stable, subject-specific neural fingerprints. Extending this approach to clinical populations could therefore enable ws-ATM–based prediction of functional decline or recovery trajectories over time, ultimately advancing toward individualized disease monitoring and prognosis.

Future studies should investigate the generalizability of the ws-ATM framework across different neurological conditions and brain states beyond rest. Additionally, this framework could be applied to examine the effects of neuromodulation techniques, such as transcranial magnetic stimulation (TMS) or deep brain stimulation (DBS), on large-scale brain dynamics.

Overall, our results show that ws-ATM provides a more stable and sample-efficient characterization of large-scale brain dynamics. Applied to resting-state MEG, ws-ATM converged faster than the original ATM, reduced inter-subject variability, and remained robust under data truncation. In line with this increased stability, ws-ATM improved the identification of ALS-related network alterations and selectively highlighted connections whose changes tracked clinical impairment, strengthening the association between pathological network reorganization and behavioural status. Together, these findings suggest that ws-ATM offers a principled and clinically informative account of neuronal avalanche propagation that could aid the development of non-invasive biomarkers for neurodegeneration, while its computational efficiency also makes it a strong candidate for real-time diagnostic use. Looking ahead, translating ws-ATM into a clinical tool task-based settings could support the design of adaptive, ws-ATM-driven closed-loop systems aimed at restoring communication and motor control as these functions progressively decline with ALS, ultimately helping preserve functional independence and quality of life.

## Ethics approval and consent to participate

The study protocol was approved by the Local Ethics Committee (University of Campania “Luigi Vanvitelli”) with protocol number 591/2018, and all participants provided written informed consent in accordance with the Declaration of Helsinki.

## Conflict of interest

The remaining authors have no conflicts of interest.

## Data and Code Availability

The study dataset has been fully collected and curated. The data supporting the findings of this study are available from the corresponding author upon reasonable request, for purpose of replication. The data are not publicly available due to the clinical nature of the cohort under study.

Code used to import data, analyse data, and generate manuscript figures are available on GitHub: https://github.com/CamiMannino/Weighted-stochastic-Avalanche-Transition-Matrix-ws-ATM-

## Data Availability

The data supporting the findings of this study are available from the corresponding author upon reasonable request, for purpose of replication. The data are not publicly available due to the clinical nature of the cohort under study.

## Acknowledgments

This work was supported by the ANR project MANET (Grant ANR-25-CE33-7747-01).

## Authors contribution

Conceptualization: C.M, P.S., M.C., and M.C.C. Methodology: C.M, P.S., M.A., M.C., and M.C.C. Investigation: C.M. Visualization: C.M. Supervision: P.S., M.C., M.C.C. Data collection and curation: M.D., F.T., P.S. Data processing: M.D., and P.S. Writing—original draft: C.M., P.S., M.C., and M.C.C. Writing—review and editing: C.M, P.S., F.T., M.C., and M.C.C.

---

**Supplementary Figure 1.**
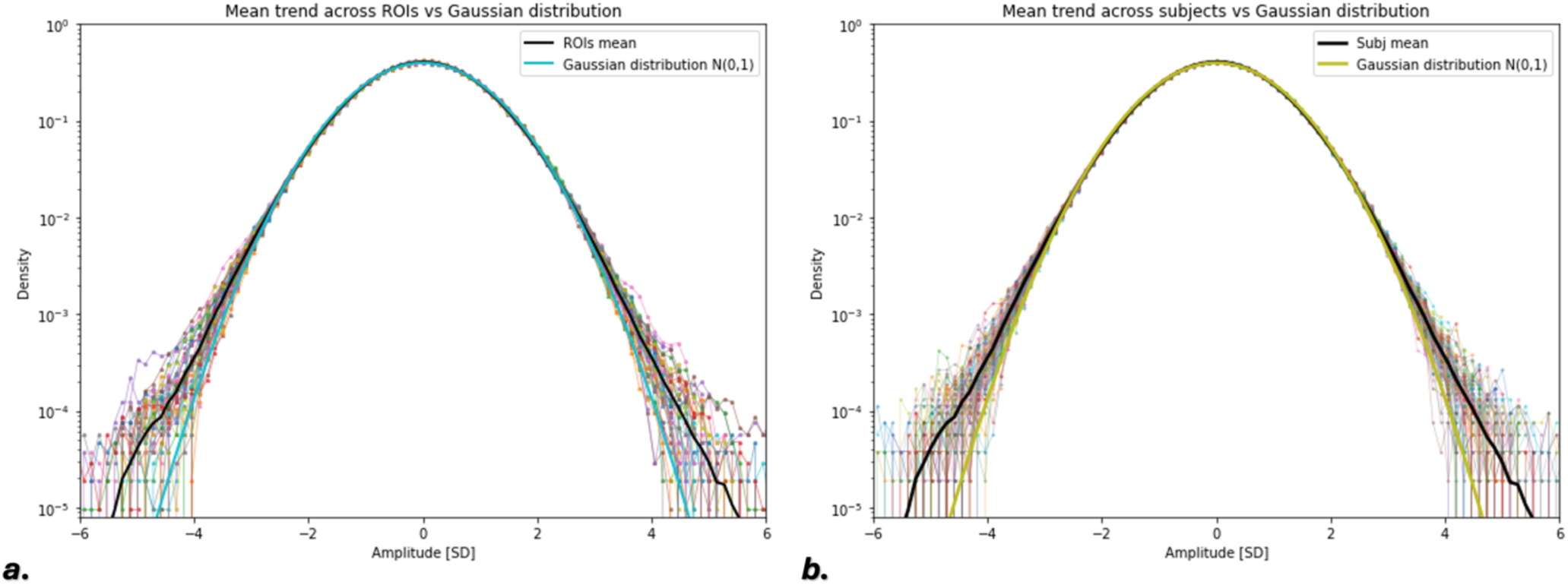
: a) Gaussian distribution vs. Neuronal Avalanches distribution over different brain regions; b) Gaussian distribution vs. Neuronal Avalanches distribution across different subjects.

**Supplementary Figure 2.**
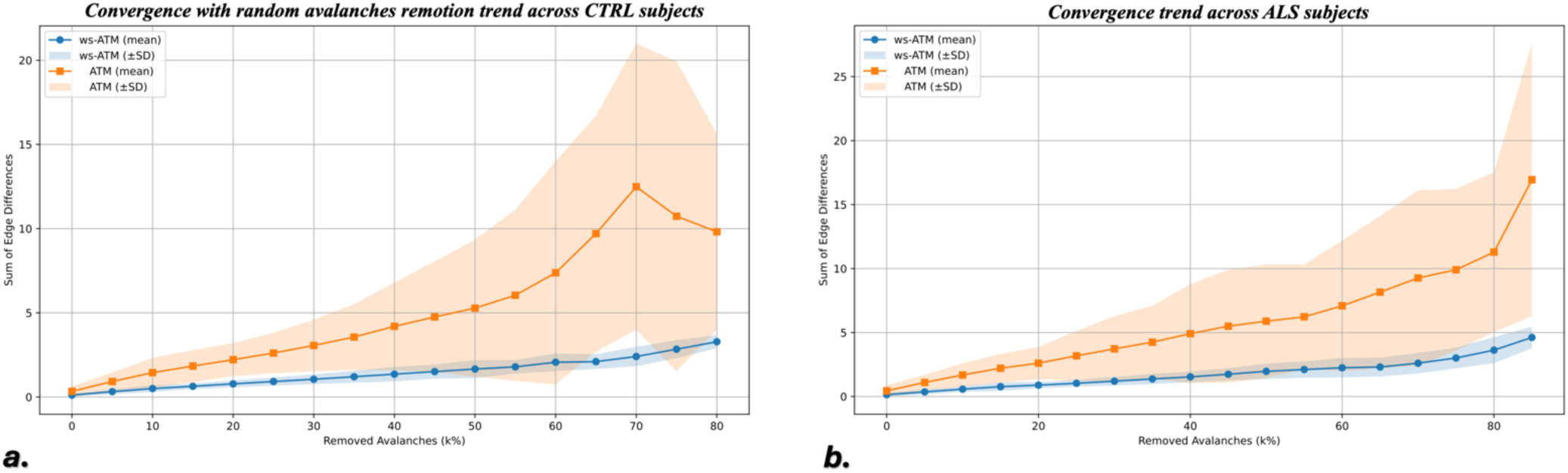
: a) Convergence trend during progressive avalanches remotion at random position in healthy subjects; b) Convergence trend during progressive avalanches remotion from the end of the signal in ALS subjects. Orange curve: Avalanches-Transition Matrix (ATM) trend; Blue curve: Weighted – Stochastic ATM (ws-ATM) trend.

**Supplementary Figure 3.**
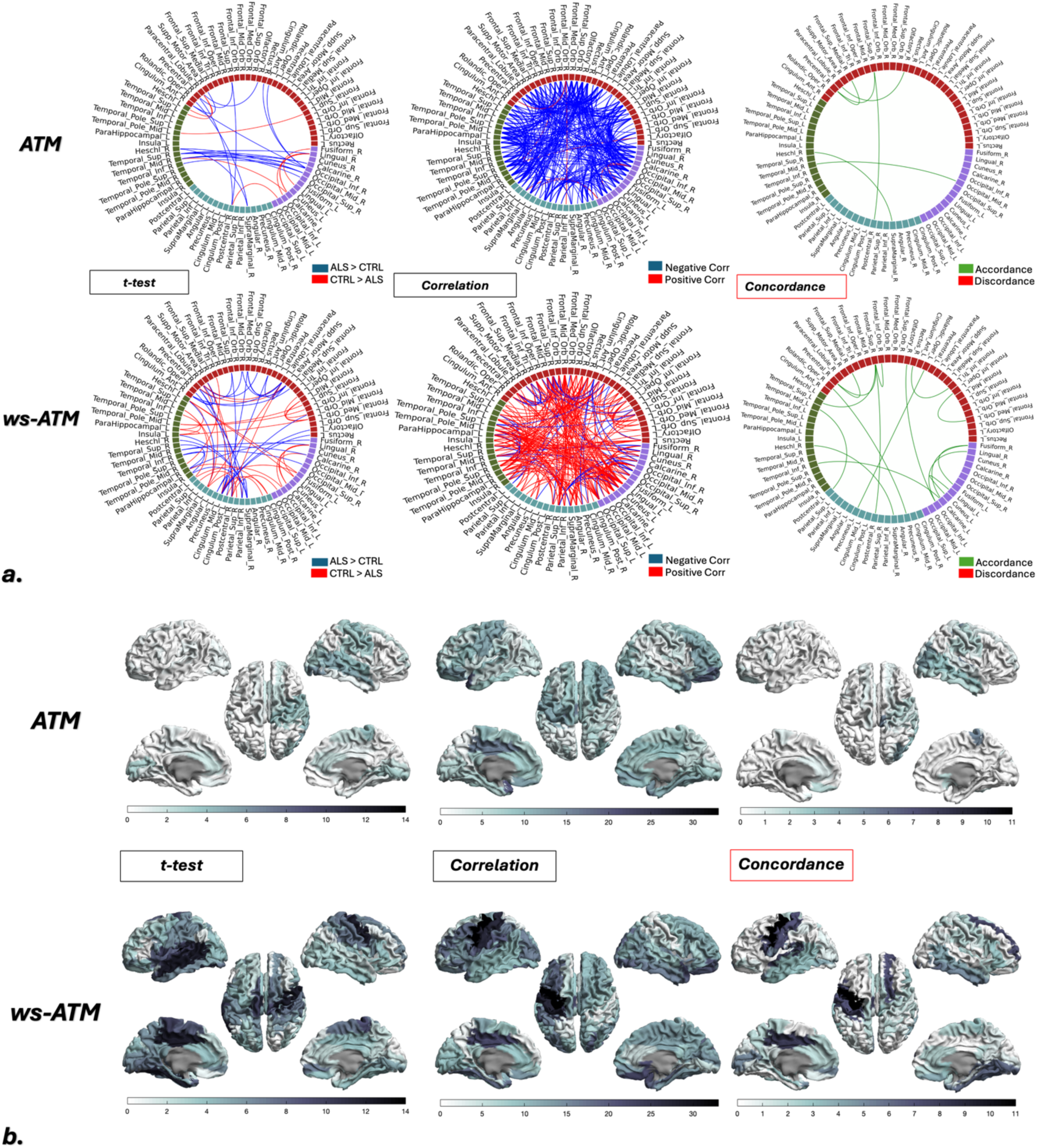
: Framework with ALSFRS-R scale: a) Edges-level, connectome : t-test analysis: ALS > CTRL Edges in blue, CTRL > ALS Edges in red; Correlation analysis: Negative correlation edges with ALSFRS-R clinical-scale in blue, Positive correlation edges with ALSFRS-R clinical-scale in red; Concordance analysis t-test & correlation : Accordance sign edges in green, Discordance sign edges in red; First row: Avalanches-Transition Matrix (ATM) Results; Second row: Weighted-Stochastic ATM (ws-ATM) Results **b) Nodal-level, brain plots:** sum of significant edges (whatever sign) for each brain region. First row: Avalanches-Transition Matrix (ATM) Results; Second row: Weighted-Stochastic ATM (ws-ATM) Results. All counts refer to edge-level tests at p < 0.05 without multiple-comparison correction.

**Supplementary Figure 4.**
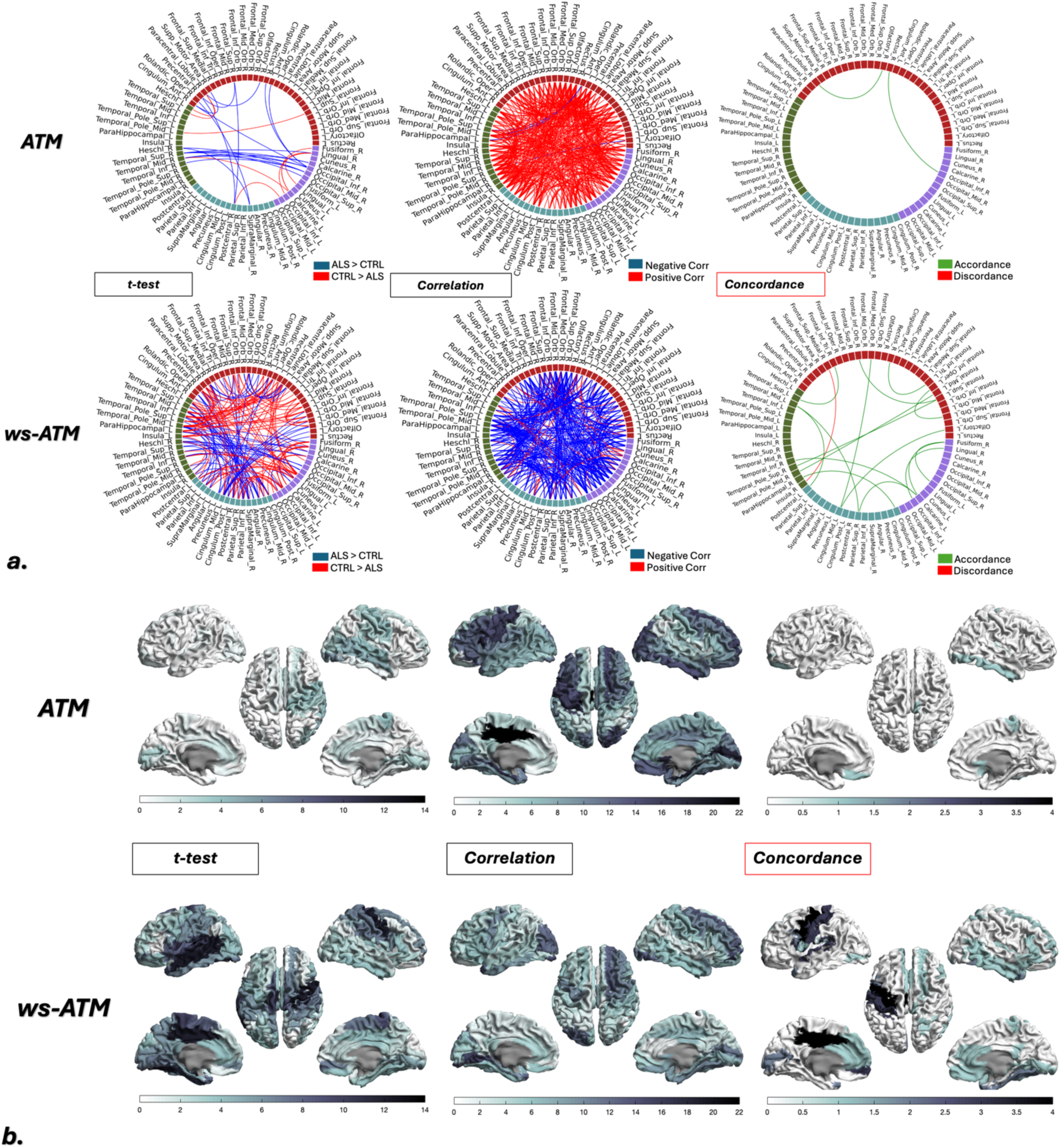
: Framework with MiToS scale: a) Edges-level, connectome : t-test analysis: ALS > CTRL Edges in blue, CTRL > ALS Edges in red; Correlation analysis: Negative correlation edges with MiTos clinical-scale in blue, Positive correlation edges with MiTos clinical-scale in red; Concordance analysis t-test & correlation : Accordance sign edges in green, Discordance sign edges in red; First row: Avalanches-Transition Matrix (ATM) Results; Second row: Weighted-Stochastic ATM (ws-ATM) Results **b) Nodal-level, brain plots:** sum of significant edges (whatever sign) for each brain region. First row: Avalanches-Transition Matrix (ATM) Results; Second row: Weighted-Stochastic ATM (ws-ATM) Results. All counts refer to edge-level tests at p < 0.05 without multiple-comparison correction.

## Notes

### Competing Interest Statement

The authors have declared no competing interest.

### Author Declarations

The study protocol was approved by the Local Ethics Committee University of Campania Luigi Vanvitelli with protocol number 591/2018, and all participants provided written informed consent in accordance with the Declaration of Helsinki.

## References

1. Rusconi M, Gerardi F, Santus W, et al. Inflammatory role of dendritic cells in Amyotrophic Lateral Sclerosis revealed by an analysis of patients’ peripheral blood. Sci Rep. 2017;7(1):7853. doi:10.1038/s41598-017-08233-1

2. Wolfson C, Gauvin DE, Ishola F, Oskoui M. Global Prevalence and Incidence of Amyotrophic Lateral Sclerosis. Neurology. 2023;101(6):e613–e623. doi:10.1212/WNL.0000000000207474

3. Cedarbaum JM, Stambler N, Malta E, et al. The ALSFRS-R: a revised ALS functional rating scale that incorporates assessments of respiratory function. J Neurol Sci. 1999;169(1):13–21. doi:10.1016/S0022-510X(99)00210-5

4. Al-Chalabi A, Chiò A, Merrill C, et al. Clinical staging in amyotrophic lateral sclerosis: analysis of Edaravone Study 19. J Neurol Neurosurg Psychiatry. 2021;92(2):165–171. doi:10.1136/jnnp-2020-323271

5. McColgan P, Joubert J, Tabrizi SJ, Rees G. The human motor cortex microcircuit: insights for neurodegenerative disease. Nat Rev Neurosci. 2020;21(8):401–415. doi:10.1038/s41583-020-0315-1

6. Verstraete E, Van Den Heuvel MP, Veldink JH, et al. Motor Network Degeneration in Amyotrophic Lateral Sclerosis: A Structural and Functional Connectivity Study. Zhan W, ed. PLoS ONE. 2010;5(10):e13664. doi:10.1371/journal.pone.0013664

7. Govaarts R, Scheijbeler EP, Beeldman E, et al. Longitudinal changes in MEG-based brain network topology of ALS patients with cognitive/behavioral impairment-An exploratory study. Netw Neurosci Camb Mass. 2025;9(3):824–841. doi:10.1162/netn_a_00450

8. Sorrentino P, Rucco R, Jacini F, et al. Brain functional networks become more connected as amyotrophic lateral sclerosis progresses: a source level magnetoencephalographic study. NeuroImage Clin. 2018;20:564–571. doi:10.1016/j.nicl.2018.08.001

9. Romano A, Trosi Lopez E, Liparoti M, et al. The progressive loss of brain network fingerprints in Amyotrophic Lateral Sclerosis predicts clinical impairment. NeuroImage Clin. 2022;35:103095. doi:10.1016/j.nicl.2022.103095

10. Beggs JM, Plenz D. Neuronal Avalanches in Neocortical Circuits. J Neurosci. 2003;23(35):11167–11177. doi:10.1523/JNEUROSCI.23-35-11167.2003

11. Haldeman C, Beggs JM. Critical Branching Captures Activity in Living Neural Networks and Maximizes the Number of Metastable States. Phys Rev Lett. 2005;94(5):058101. doi:10.1103/PhysRevLett.94.058101

12. Shriki O, Alstott J, Carver F, et al. Neuronal Avalanches in the Resting MEG of the Human Brain. J Neurosci. 2013;33(16):7079–7090. doi:10.1523/JNEUROSCI.4286-12.2013

13. Sorrentino P, Seguin C, Rucco R, et al. The structural connectome constrains fast brain dynamics. eLife. 2021;10:e67400. doi:10.7554/eLife.67400

14. Tewarie P, Prasse B, Meier J, et al. Predicting time-resolved electrophysiological brain networks from structural eigenmodes. Hum Brain Mapp. 2022;43(14):4475–4491. doi:10.1002/hbm.25967

15. Duma GM, Danieli A, Mento G, et al. Altered spreading of neuronal avalanches in temporal lobe epilepsy relates to cognitive performance: A resting-state hdEEG study. Epilepsia. 2023;64(5):1278–1288. doi:10.1111/epi.17551

16. Corsi MC, Troisi Lopez E, Sorrentino P, et al. Neuronal avalanches in temporal lobe epilepsy as a noninvasive diagnostic tool investigating large scale brain dynamics. Sci Rep. 2024;14:14039. doi:10.1038/s41598-024-64870-3

17. Mannino C, Sorrentino P, Chavez M, Corsi MC. Neuronal avalanches as a predictive biomarker for guiding tailored BCI training programs. bioRxiv. Preprint posted online August 12, 2025:2025.05.31.657206. doi:10.1101/2025.05.31.657206

18. Polverino A, Troisi Lopez E, Minino R, et al. Flexibility of Fast Brain Dynamics and Disease Severity in Amyotrophic Lateral Sclerosis. Neurology. 2022;99(21):e2395–e2405. doi:10.1212/WNL.0000000000201200

19. Polverino A, Troisi Lopez E, Liparoti M, et al. Altered spreading of fast aperiodic brain waves relates to disease duration in Amyotrophic Lateral Sclerosis. Clin Neurophysiol. 2024;163:14–21. doi:10.1016/j.clinph.2024.04.003

20. Corsi MC, Sorrentino P, Schwartz D, et al. Measuring neuronal avalanches to inform brain-computer interfaces. iScience. 2024;27(1):108734. doi:10.1016/j.isci.2023.108734

21. Mannino, Camilla, Sorrentino, Pierpaolo, Chavez, Mario, Corsi, Marie-Constance. Neuronal Avalanches for EEG-based Motor Imagery BCI. 2024;GBCIC 2024-9th Graz Brain-Computer Interface Conference 2024:98–104. doi:10.3217/978-3-99161-014-4-018

22. Brooks BR. El escorial World Federation of Neurology criteria for the diagnosis of amyotrophic lateral sclerosis. J Neurol Sci. 1994;124:96–107. doi:10.1016/0022-510X(94)90191-0

23. Oostenveld R, Fries P, Maris E, Schoffelen JM. FieldTrip: Open Source Software for Advanced Analysis of MEG, EEG, and Invasive Electrophysiological Data. Comput Intell Neurosci. 2011;2011:156869. doi:10.1155/2011/156869

24. Gong G, He Y, Concha L, et al. Mapping Anatomical Connectivity Patterns of Human Cerebral Cortex Using In Vivo Diffusion Tensor Imaging Tractography. Cereb Cortex. 2009;19(3):524–536. doi:10.1093/cercor/bhn102

25. Attal Y, Schwartz D. Assessment of Subcortical Source Localization Using Deep Brain Activity Imaging Model with Minimum Norm Operators: A MEG Study. PLOS ONE. 2013;8(3):e59856. doi:10.1371/journal.pone.0059856

26. de Pasquale F, Della Penna S, Snyder AZ, et al. Temporal dynamics of spontaneous MEG activity in brain networks. Proc Natl Acad Sci U S A. 2010;107(13):6040–6045. doi:10.1073/pnas.0913863107

27. Norris JR, ed. Continuous-time Markov chains I. In: Markov Chains. Cambridge Series in Statistical and Probabilistic Mathematics. Cambridge University Press; 1997:60–107. doi:10.1017/CBO9780511810633.004

28. Norris JR, ed. Continuous-time Markov chains II. In: Markov Chains. Cambridge Series in Statistical and Probabilistic Mathematics. Cambridge University Press; 1997:108–127. doi:10.1017/CBO9780511810633.005

29. Stam CJ. Modern network science of neurological disorders. Nat Rev Neurosci. 2014;15(10):683–695. doi:10.1038/nrn3801

30. Zalesky A, Fornito A, Bullmore ET. Network-based statistic: Identifying differences in brain networks. NeuroImage. 2010;53(4):1197–1207. doi:10.1016/j.neuroimage.2010.06.041

31. Cipriano L, Minino R, Liparoti M, et al. Flexibility of brain dynamics is increased and predicts clinical impairment in relapsing-remitting but not in secondary progressive multiple sclerosis. Brain Commun. 2024;6(2):fcae112. doi:10.1093/braincomms/fcae112

32. Romano A, Troisi Lopez E, Cipriano L, et al. Topological changes of fast large-scale brain dynamics in mild cognitive impairment predict early memory impairment: a resting-state, source reconstructed, magnetoencephalography study. Neurobiol Aging. 2023;132:36–46. doi:10.1016/j.neurobiolaging.2023.08.003

33. Agosta F, Valsasina P, Absinta M, et al. Sensorimotor Functional Connectivity Changes in Amyotrophic Lateral Sclerosis. Cereb Cortex. 2011;21(10):2291–2298. doi:10.1093/cercor/bhr002

34. Schmidt R, Verstraete E, de Reus MA, Veldink JH, van den Berg LH, van den Heuvel MP. Correlation between structural and functional connectivity impairment in amyotrophic lateral sclerosis. Hum Brain Mapp. 2014;35(9):4386–4395. doi:10.1002/hbm.22481

35. Arviv O, Goldstein A, Shriki O. Neuronal avalanches and time-frequency representations in stimulus-evoked activity. Sci Rep. 2019;9(1):13319. doi:10.1038/s41598-019-49788-5

36. Palva JM, Zhigalov A, Hirvonen J, Korhonen O, Linkenkaer-Hansen K, Palva S. Neuronal long-range temporal correlations and avalanche dynamics are correlated with behavioral scaling laws. Proc Natl Acad Sci U S A. 2013;110(9):3585–3590. doi:10.1073/pnas.1216855110

37. Trubshaw M, Gohil C, Yoganathan K, et al. The cortical neurophysiological signature of amyotrophic lateral sclerosis. Brain Commun. 2024;6(3):fcae164. doi:10.1093/braincomms/fcae164

38. Agosta F, Sala S, Valsasina P, et al. Brain network connectivity assessed using graph theory in frontotemporal dementia. Neurology. 2013;81(2):134–143. doi:10.1212/WNL.0b013e31829a33f8

39. Zhou F, Gong H, Li F, et al. Altered motor network functional connectivity in amyotrophic lateral sclerosis: a resting-state functional magnetic resonance imaging study. NeuroReport. 2013;24(12):657. doi:10.1097/WNR.0b013e328363148c

40. Devine MS, Pannek K, Coulthard A, McCombe PA, Rose SE, Henderson RD. Exposing asymmetric gray matter vulnerability in amyotrophic lateral sclerosis. NeuroImage Clin. 2015;7:782–787. doi:10.1016/j.nicl.2015.03.006

41. Precentral degeneration and cerebellar compensation in amyotrophic lateral sclerosis: A multimodal MRI analysis. doi:10.1002/hbm.24609

42. Li F, Zhou F, Huang M, Gong H, Xu R. Frequency-Specific Abnormalities of Intrinsic Functional Connectivity Strength among Patients with Amyotrophic Lateral Sclerosis: A Resting-State fMRI Study. Front Aging Neurosci. 2017;9. doi:10.3389/fnagi.2017.00351

43. Precentral degeneration and cerebellar compensation in amyotrophic lateral sclerosis: A multimodal MRI analysis. doi:10.1002/hbm.24609

44. Trojsi F, Monsurrò MR, Esposito F, Tedeschi G. Widespread Structural and Functional Connectivity Changes in Amyotrophic Lateral Sclerosis: Insights from Advanced Neuroimaging Research. Neural Plast. 2012;2012:473538. doi:10.1155/2012/473538

45. Goldstein LH, Abrahams S. Changes in cognition and behaviour in amyotrophic lateral sclerosis: nature of impairment and implications for assessment. Lancet Neurol. 2013;12(4):368–380. doi:10.1016/S1474-4422(13)70026-7

46. Trojsi F, Sorrentino P, Sorrentino G, Tedeschi G. Neurodegeneration of brain networks in the amyotrophic lateral sclerosis–frontotemporal lobar degeneration (ALS–FTLD) continuum: evidence from MRI and MEG studies. CNS Spectr. 2018;23(6):378–387. doi:10.1017/S109285291700075X

47. Beeldman E, Govaarts R, de Visser M, et al. Screening for cognition in amyotrophic lateral sclerosis: test characteristics of a new screen. J Neurol. 2021;268(7):2533–2540. doi:10.1007/s00415-021-10423-x

48. Voxel-based morphometry study of brain volumetry and diffusivity in amyotrophic lateral sclerosis patients with mild disability.doi:10.1002/hbm.20364

49. Bede P, Hardiman O. Longitudinal structural changes in ALS: a three time-point imaging study of white and gray matter degeneration. Amyotroph Lateral Scler Front Degener. 2018;19(3-4):232–241. doi:10.1080/21678421.2017.1407795

50. Borgheai SB, McLinden J, Mankodiya K, Shahriari Y. Frontal Functional Network Disruption Associated with Amyotrophic Lateral Sclerosis: An fNIRS-Based Minimum Spanning Tree Analysis. Front Neurosci. 2020;14. doi:10.3389/fnins.2020.613990

51. Ahmed RM, Devenney EM, Irish M, et al. Neuronal network disintegration: common pathways linking neurodegenerative diseases. J Neurol Neurosurg Psychiatry. 2016;87(11):1234–1241. doi:10.1136/jnnp-2014-308350

52. Warren JD, Rohrer JD, Schott JM, Fox NC, Hardy J, Rossor MN. Molecular nexopathies: a new paradigm of neurodegenerative disease. Trends Neurosci. 2013;36(10):561–569. doi:10.1016/j.tins.2013.06.007

53. Bede P. Deciphering neurodegeneration. *Neurology*. 2017;89(17):1758-1759. doi:10.1212/WNL.0000000000004582

54. Bede P, Omer T, Finegan E, et al. Connectivity-based characterisation of subcortical grey matter pathology in frontotemporal dementia and ALS: a multimodal neuroimaging study. Brain Imaging Behav. 2018;12(6):1696–1707. doi:10.1007/s11682-018-9837-9

55. Hardiman O, Al-Chalabi A, Chio A, et al. Amyotrophic lateral sclerosis. Nat Rev Dis Primer. 2017;3(1):17071. doi:10.1038/nrdp.2017.71

56. Verde F, Del Tredici K, Braak H, Ludolph A. The multisystem degeneration amyotrophic lateral sclerosis - neuropathological staging and clinical translation. Arch Ital Biol. 2017;155(4):118–130. doi:10.12871/00039829201746

